# Highly performing point-of-care molecular testing for SARS-CoV-2 with RNA extraction and isothermal amplification

**DOI:** 10.1101/2020.08.01.20166538

**Authors:** Pierre Garneret, Etienne Coz, Elian Martin, Jean-Claude Manuguerra, Elodie Brient-Litzler, Vincent Enouf, Daniel Felipe González Obando, Jean-Christophe Olivo-Marin, Fabrice Monti, Sylvie Van der Werf, Jessica Vanhomwegen, Patrick Tabeling

## Abstract

In order to respond to the urgent request of massive testing, developed countries perform nucleic acid amplification tests (NAAT) of SARS-CoV-2 in centralized laboratories. Real-time RT - PCR (Reverse transcription - Polymerase Chain Reaction) is used to amplify the viral RNA and enable its detection. Although PCR is 37 years old, it is still considered, without dispute, as the gold standard. PCR is an efficient process, but the complex engineering required for automated RNA extraction and temperature cycling makes it incompatible for use in point of care settings. In the present work, by harnessing progress made in the past two decades in DNA amplification, microfluidics and membrane technologies, we succeeded to create a portable test, in which SARS-CoV-2 RNA is extracted, amplified isothermally by RT - LAMP (Loop-mediated Isothermal Amplification), and detected using intercalating dyes or highly fluorescent probes. Depending on the viral load, the detection takes between twenty minutes and one hour. Using pools of naso-pharyngal clinical samples, we estimated a sensitivity comparable to RT-qPCR (up to a Cycle threshold of 39, equivalent to <0.1 TCID50 per mL) and a 100% specificity, for other human coronaviruses and eight respiratory viruses currently circulating in Europe. We designed and fabricated an easy-to-use portable device called “COVIDISC” to carry out the test at the point of care. The low cost of the materials along with the absence of complex equipment paves the way towards a large dissemination of this device. The perspective of a reliable SARS-CoV-2 point of care detection, highly performing, that would deliver on-site results in less than one hour, with a self-testing potential, opens up a new efficient approach to manage the pandemics.

## Introduction

Nucleic Acid Amplification Tests (NAATs) detect, through amplification of the nucleic acids, the presence of pathogens in infected samples. They are characterized by high sensitivities (10-100 viral particles) and excellent specificities. There are different ways of performing NAATs but today, PCR (Polymerase Chain Reaction), coupled to extraction, is considered as the gold standard.

The main problem of PCR is that, due to an inherent complexity, the only way to obtain high throughputs is to perform highly parallelized tests in centralized laboratories. This generates logistics issues, long delivery times, (several days in practice) and costs unaffordable for developing countries. Over the last ten years, with the advent of isothermal amplification technologies *(2)*, a new area of research has opened, raising hope to perform NAATs at the point of care, with portable devices much cheaper than extraction/PCR platforms and showing comparable performances. A new generation of NAATs has grown along the years, in laboratories, often coupling isothermal amplification to paper microfluidics, a field pioneered by G.Whitesides *(3-16)*.

Unfortunately, this second generation of testing, despite its potential, did not succeed to take off as a point of care (POC) product, for several reasons (low practicability of the devices, absence of sample preparation, in particular RNA extraction, clinical and analytical performances still to be assessed, costly surrounding equipment, competitiveness,…). Thereby, as SARS-CoV-2 started to propagate around the world, the isothermal technology was not ready to face the urgent request of global massive testing. Over these last weeks, stimulated by the crisis, isothermal commercial machines have been proposed *(17)*. However, because of cost, throughput and performance limitations, they do not represent yet an alternative to the current extraction/RT-PCR technology. Laboratory isothermal detection of SARS-CoV-2 have been reported in the literature, but only in microtube strips *(18-21)*.

By carrying out series of improvements and optimizations over the last six years, we made a technological leap in the domain. The new generation of molecular tests we report here combines membrane extraction, RT-LAMP testing *(22)* and paper microfluidics. It has performances comparable to the extraction/RT-PCR method, with a potential to be used promptly for the detection of SARS-CoV-2 at the point of care.

## Materials and methods

The laboratory device we used is displayed on Fig 1A. Two sheets of polypropylene (black), two centimeters in size, incorporate pretreated laser-cut disks, one (oblong form) for nucleic acid extraction, and the two others (circular disks) for the RT-LAMP reactions. The oblong disk is a commercial silicon membrane similar to those optimized for RNA extraction (23), the others are made of glass fiber (Watman). Figure 1B decomposes the workflow. The lysed sample is injected onto the extraction membrane and washed *(24-25)*. After drying, the device is folded so as to place the oblong extraction membrane in contact withthe two reaction disks. Then RNA elution is achieved, using the rehydrationbuffer. In this process, the captured RNA is driven towards the reaction disk, by capillary action. On both disks, the reaction mix gets hydrated and, upon temperature rise to 65°C, reverse transcription and amplification are performed. We use real-time RT-LAMP, whose kinetics is measured by tracking the fluorescence emission of a DNA intercalating dye.

**Fig. 1:**
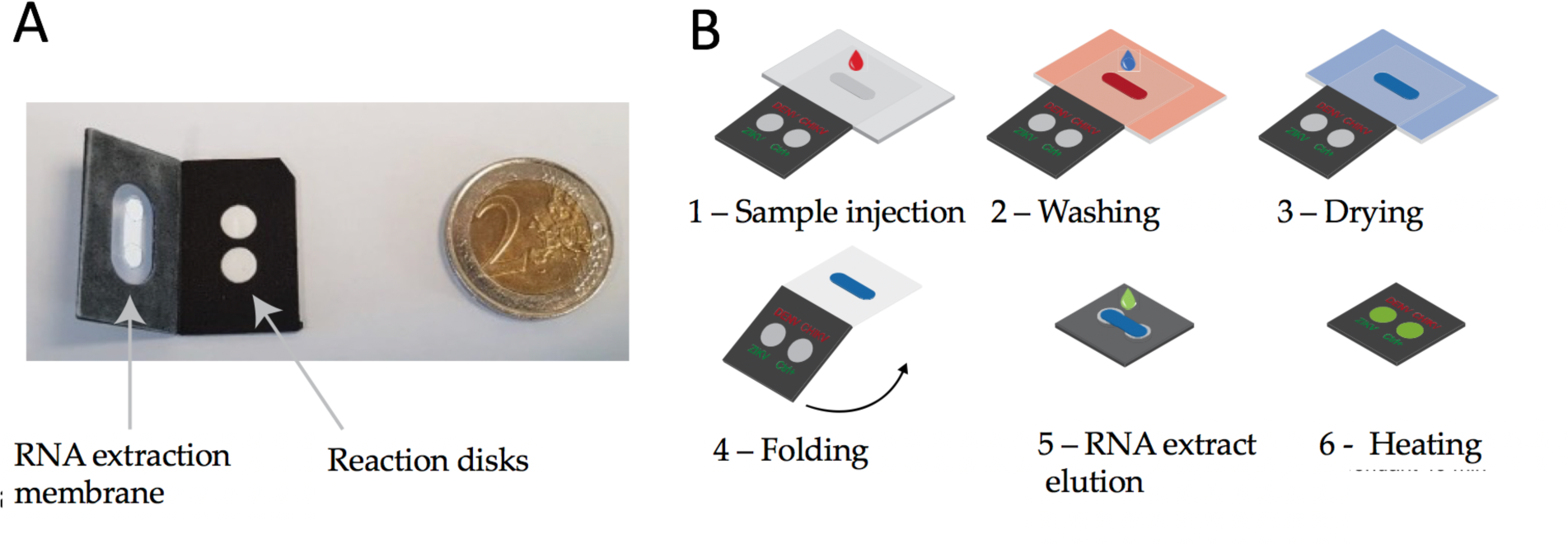
Laboratory device and workflow. A: Two sheets of polypropylene (black) in which pretreated pieces of glass fiber (in white) are incorporated. B: displays the mode of operation. 1 – The sample, in which the virus has been lysed, is injected onto the extraction membrane. 2 – Washing. 3 – Drying of the extraction membrane. 4 – Folding of the device in such a way that the extraction membrane comes into contact with the two reaction disks. On both disk is freeze-dried the LAMP mix and primers (Covid-19 and human 18S RNA for the test and control disk respectively) permiting reverse transcription and amplification. 5 – Elution of the RNA from the capture membrane to the reaction disks, then, sealing with PCR tape 6 – Heating at 65°C. Read-out in real time with an intercalating agent (SYTO82).

## Results

We now use the device of Fig. 1 for the detection of SARS-CoV-2. The primers are displayed in Supp Mat 1 *(19)*. Figure 2 shows end point results, sixty minutes after sample injection. We used eight pools of naso-pharyngal samples infected with different SARS-CoV-2 viral loads, in duplicate. All were characterized with the real-time duplex RT-PCR (targeting the RdRP gene) developed by CNR Pasteur *(26)*. In terms of cycle thresholds (Ct), they span a range extending from 15 to 39, i.e down to a few viral particles per milliliter *(27)*. For the specificity, we used clinical naso-pharyngal samples infected by respiratory pathogens: Respiratory syncytial virus (RSV) A and B, influenza A(H1) virus and influenza A(H3) virus, influenza B Victoria (BVIC) and Yamagata lineages (BYAM), human rhinovirus (HRV) and human metapneumovirus (hMPV). In addition, testing of high-titer solutions of human coronaviruses in microtubes, including SARS-CoV, MERS-CoV, OC43 and 229E demonstrated a 100% specificity towards these closely-related viruses.

**Fig 2.**
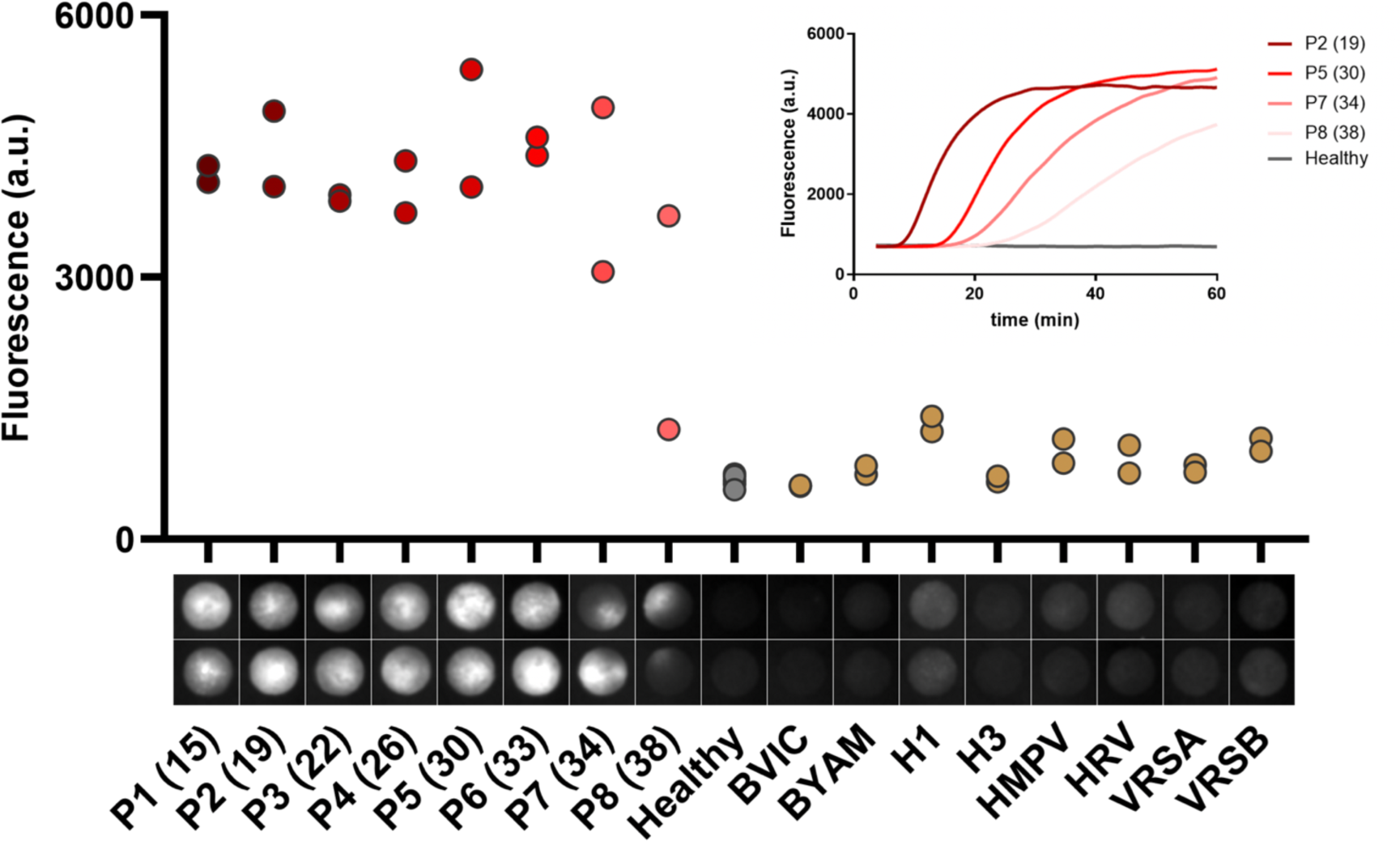
Sensitivity and specificity of the test: Sensitivity and Specificity end point (t=60 min) measurements obtained on SARS-CoV-2 positive samples P1-P8 (Cts in parenthesis), negative samples (individuals with a negative RT-PCR)and, on the right part, patients diagnosed with other respiratory infections. The low intensity level measured on the negative samples (grey dots) corresponds to background fluorescence. The internal controls (RNA 18s) are displayed in Supp Mat 2. At the bottom, series of disk images obtained at the end point, each vertical pair corresponding to duplicate assays. (Insert) RT-LAMP amplification curves obtained by real-time monitoring of the fluorescence produced by an intercalating dye (SYTO82).

Below the abscissa of Fig. 2, we show the fluorescence images of the reaction disks corresponding to the end point measurements. Heterogeneities of the fluorescence field are visible on most of them, a phenomenon we attribute, speculatively, to heterogeneous nucleation and low diffusion of the amplicons inside the porous medium. The fluorescence signal plotted on Figure 2 is the average value of the 25% brighter pixels within the reaction disk, a simple approach to take into account profiles of low Ct samples, where the reaction is confined in a subpart of the disk. This feature suggests that a classification based on the fluorescence standard deviation could also be envisaged (see SupMat 4).

Fig. 2 shows that we clearly detect patient samples 1-7, i.e up to a cycle threshold of 35. The sample n°8 at Ct=39 is also detected, but with a substantial dispersion. We conclude that the limit-of-detection of the method is equivalent to the extraction/RT-PCR performances.

The insert of Fig 2 shows some kinetics of the reactions, for different viral loads, using a fluorescent intercalating agent (SYTO82) *(28)*. The negative control remains at a level corresponding to the natural fluorescence of the reaction disk. Amplification curves have the usual sigmoid shape. The sample with the highest viral load, (Ct=19, equivalent to an infectious titer of 10^5^ TCID50 per mL of sample), can be detected after 15 minutes. At higher Ct values, i.e. as viral load decreases, the signal takes off at later times. Plateaus are observed for the patients 1-6, i.e up to Ct=33. For the samples with the lowest viral loads (pools 7 and 8), i.e., 100-10 genome copies per reaction respectively, a plateau is not reached, but still, the virus can be detected without ambiguity.

The right part of Figure 2 addresses the question of the specificity. The images show that for non-SARS-CoV-2 pathogens, fluorescence is barely visible. This is confirmed by intensity measurements, which show that the fluorescence level is indistinguishable from the background. One may conclude that the specificity of the test, based on this limited set of pathogens, is 100%.

In order to transform the foldable paper system into a practical POC device, we created the “COVIDISC”, shown in Fig 3.

**Figure 3:**
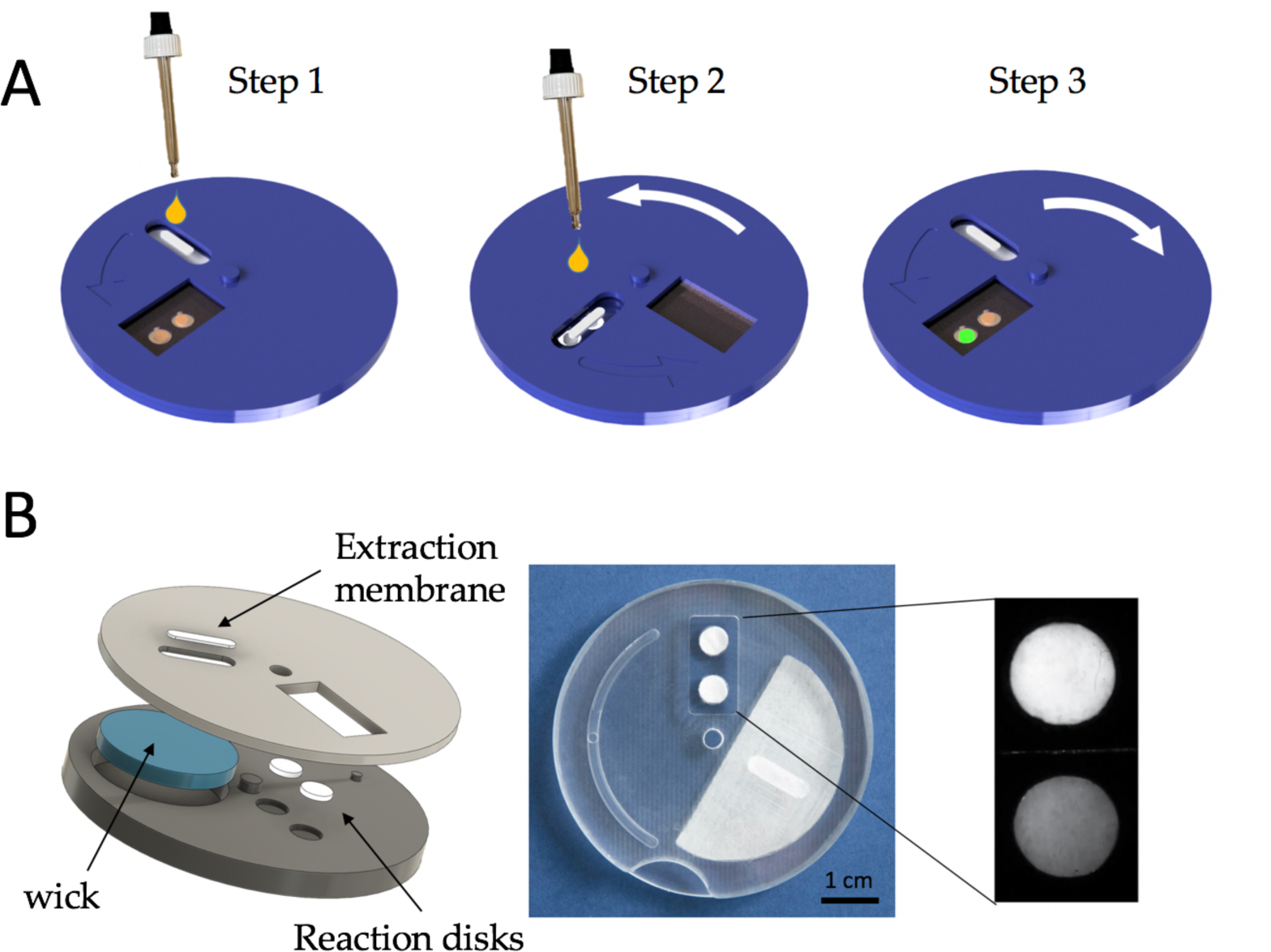
Description of the COVIDISC. A – COVIDISC workflow decomposed in three steps: 1 - injection, washing, drying. 2 - Disk rotation and elution; 3 – Disk counter-rotation, coverage of the reaction zone by a PCR sealing film, heating, amplification and readout. B – Left: Exploded structure of the device. Center: Picture of a prototype. Right: QUASR readout photograph of a test on RNA extracts of SARS-CoV-2, processed as in Fig 3A; (left) Positive sample; (right) negative sample.

Fig 3A shows the COVIDISC workflow, which reproduces that of Fig 1B. The device, shown in Fig 3B consists of two plastic disks, 5 cm in diameter, able to rotate around a common axis. The extraction membrane, the wick and the reaction disks are force fitted. By performing rotations, injections and heating, one executes the workflow of Fig 1B (see SuppMat 4). On adding QUASR probes (Quenching of Unincorporated Amplification Signal Reporters) (29) to RT-LAMP, we obtain a naked eye YES/NO answer. Fig 3B (right) shows the readout of RNA extracts of SARS-CoV-2, captured with the camera of a smart phone. We used an oven to maintain the temperature at 65°C and one LED and two gelatin filters to take the picture. The equipment needed for running the test is thus minimal.

## Conclusion

The results presented here have been obtained with 39 clinical samples. Although a confirmation on larger sets is desirable, our measurements indicate that, with a minimal equipment, one can extract, wash, elute, reverse-transcribe, amplify and measure the kinetics, with a sensitivity comparable to the gold standard RT-qPCR, i.e. 39 cycles (a few genome copies per reaction) and a specificity, based on the set of pathogens we used, of 100%. We also created a portable device (COVIDISC) that can be used at the point of care with minimal equipment *(30)*. This work paves the way toward reliable point of care testing of SARS-CoV-2, both in developing and developed countries. Performing these tests at the doctor’s office, at the working place or in pharmacies could allow the isolation of infected patients without delay, fastenning their quarantine,reducing logistics and costs and offering a new efficient approach to manage the pandemics. In the future, obviously, the same technology could be used for other pathogens.

## Data Availability

All data is in the paper

## Acknowlegment

The authors thank ESPCI, CNRS, IPGG platform (ANR-10-EQPX-34), PSL, PSL-VALO, PASTEUR, Carnot Pasteur Maladies Infectieuses program (ANR 11-CARN 017-01), PARIS City, ANR Flash, DIM ELICIT, Fonds de l’ESPCI, Bettencourt Fondation for their support. They thank DNA SCRIPT, France Ambassador in JAPAN, M.Ban, Bertin-Technology, Withings, T.Coulhon, JL.Missika, MC.Lemardeley, V.Croquette, A.Boquet-Pujadas, C.Batéjat, D.Hoinard, L.Dehove, A.Kwasiborski, Q.Grassin, M.Feher, for discussion and help

## Ethical documentation

Samples used in this study were collected as part of approved ongoing surveillance conducted by the National Reference Center for Respiratory Viruses (NRC) at Institut Pasteur (WHO reference laboratory providing confirmatory testing for COVID-19). The lab investigations described in this article were carried out in accordance with French Law which does not required a review by an ethics committee for the secondary use of samples collected for healthcare purposes. However, to fulfill editorial requirements, an Ethics waiver (reference 2020/08) has been obtained by the Institut Pasteur from a French Ethics Committee (Comité de Protection des Personnes Ile de France IV, IRB#00003835).

## SUPPLEMENTARY MATERIALS

### SUPPLEMENTARY MATERIAL 1

#### Sequences of the primers used in the SARS-CoV2 test (From Lamb et al (19,28))

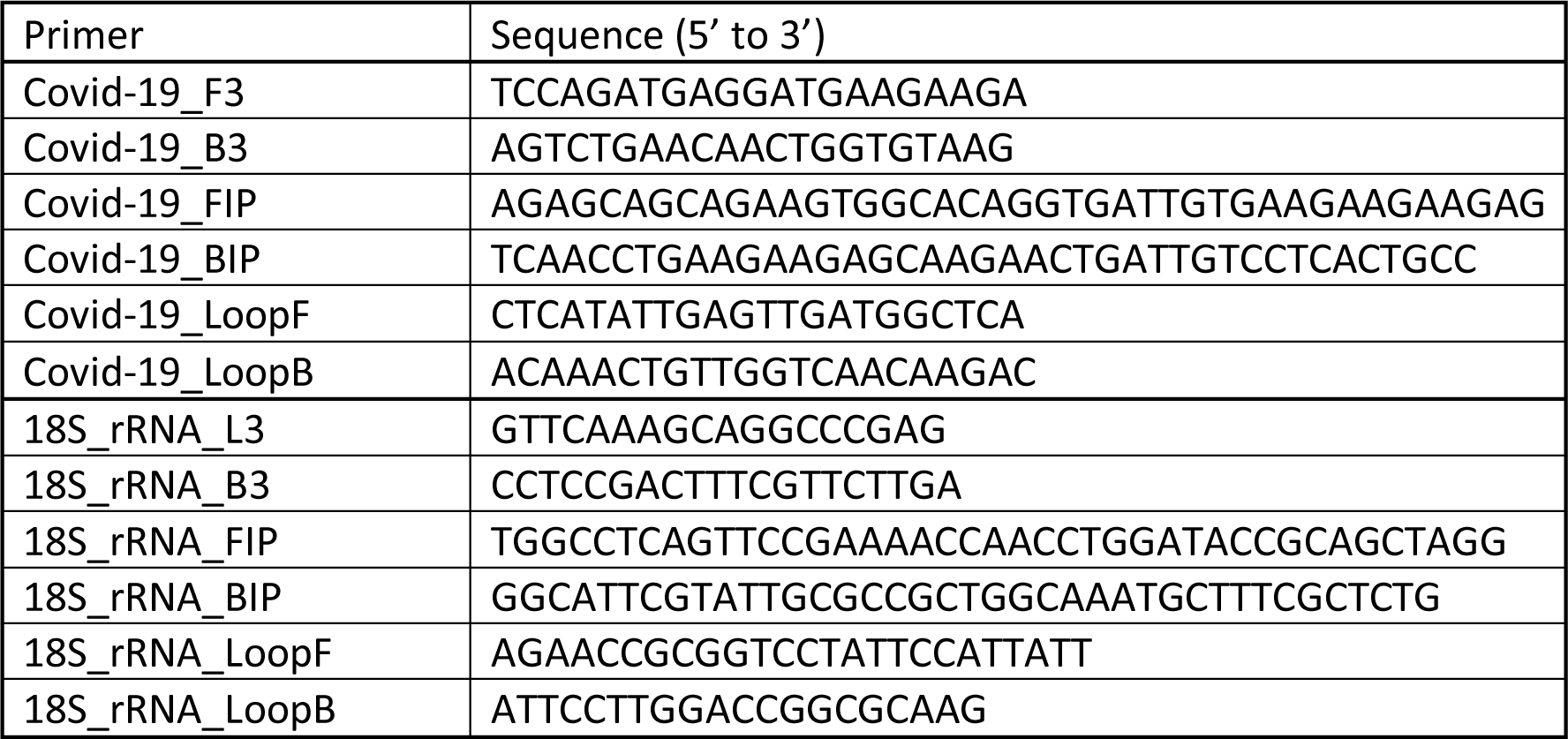

### SUPPLEMENTARY MATERIAL 2

#### Positive controls analysis

In our device, each clinical sample is split into two reaction disks: a SARS-CoV-2 test and an internal control (human 18S ribosomal RNA) test. This internal control can help identify several types of problems: (i) too little human material in the sample; (ii) the RNA extraction step did not work; (iii) the reagents lyophilized on the device don’t work anymore (e.g. due to wrong storage conditions, expired device…); (iv) the amplification did not work due to a misfunction of the heater. The result of the SARS-CoV-2 test should be rejected if the internal control test is not positive.

For each data point presented in Figure 2, we ran an internal control on the same device, with a RT-LAMP mix detecting human 18S ribosomal RNA. The histogram of these 39 data points is represented on Figure S3. The minimal signal of all these controls (2312 a.u.) is well above the average value of the no template controls (703 a.u. ± 52 a.u, 5 replicates). (standard deviation)) (See Fig S3). The histogram is substantially broad; it may reflect the fact that the amount of human material, and thereby that of 18S RNA, may vary from one sample to the other.

**Fig. S2:**
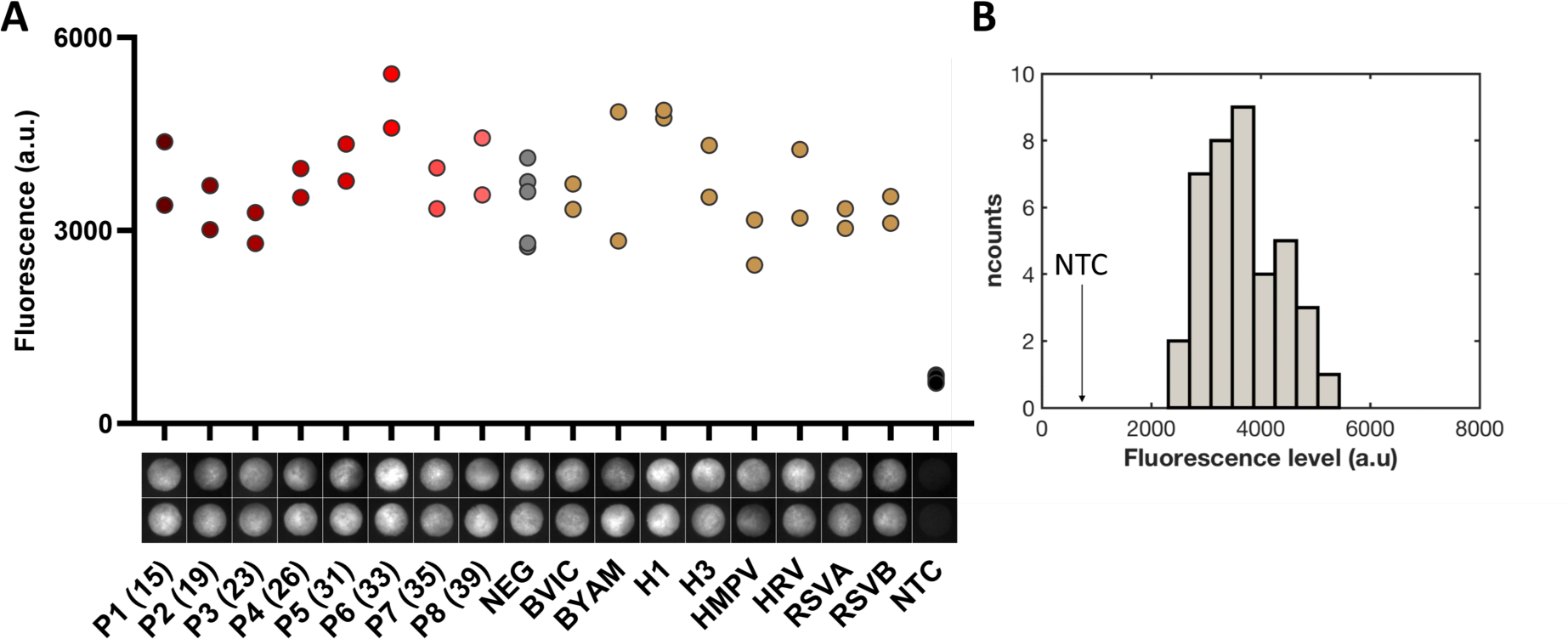
A – Human 18S RNA internal control on naso-pharyngal samples: end point (t=60 min) measurements of each internal control disks respectively corresponding to Fig.2 tests disks. Black dots display no template controls (NTC), 5 replicates. B – Histogram of the fluorescence gray levels of the 39 human 18S RNA internal controls, with a class interval of 390 units. Each level is the averaged intensity of the 25% brighter pixels of the end point image. The average fluorescence level of the no template controls (NTC) located at a level of 703 is shown on the graph.

### SUPPLEMENTARY MATERIAL 3

#### List of operations to be realized to perform the test on the COVIDISC

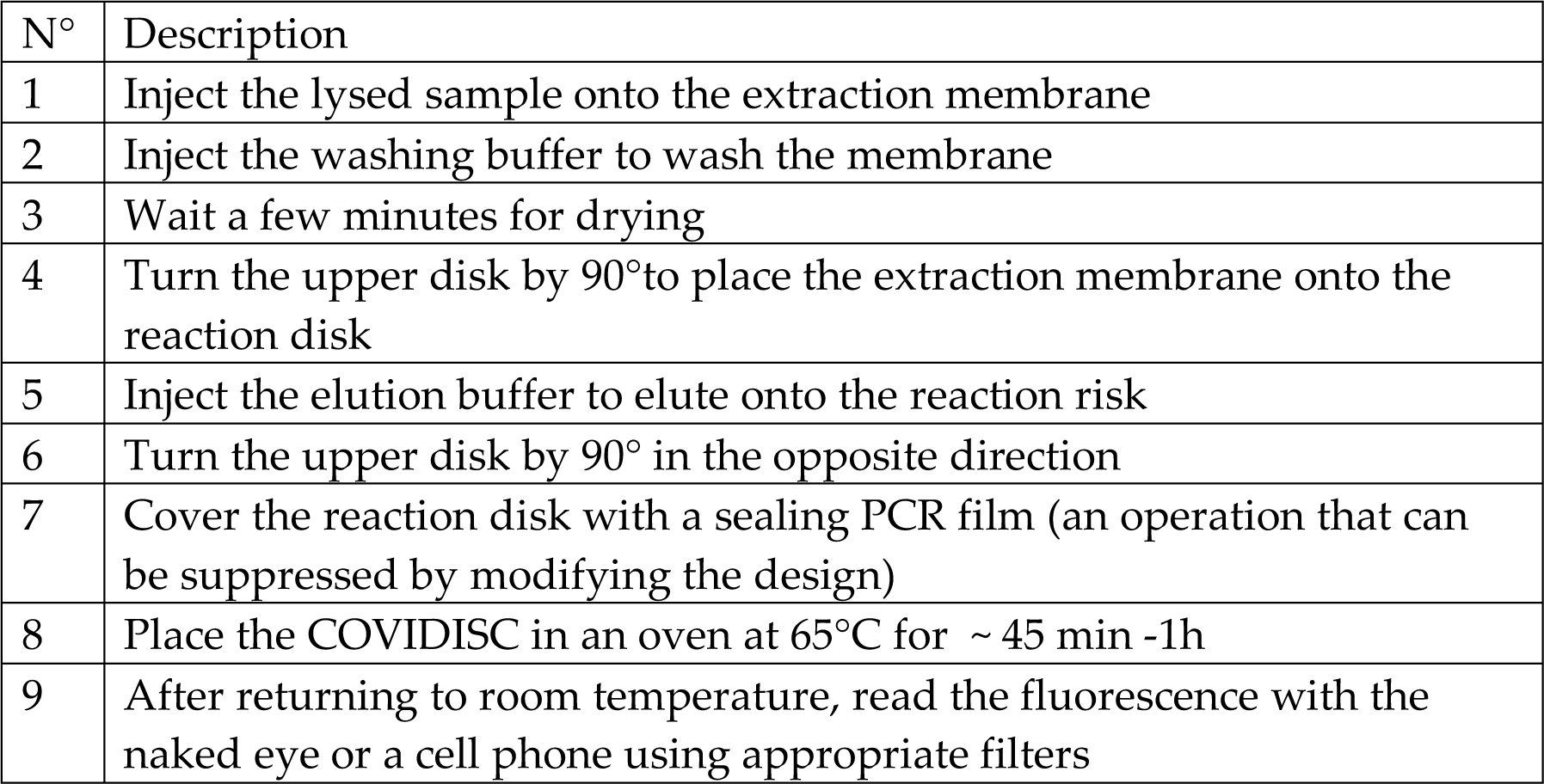

| N° | Description |
| --- | --- |
| 1 | Inject the lysed sample onto the extraction membrane |
| 2 | Inject the washing buffer to wash the membrane |
| 3 | Wait a few minutes for drying |
| 4 | Turn the upper disk by 90° to place the extraction membrane onto the reaction disk |
| 5 | Inject the elution buffer to elute onto the reaction risk |
| 6 | Turn the upper disk by 90° in the opposite direction |
| 7 | Cover the reaction disk with a sealing PCR film (an operation that can be suppressed by modifying the design) |
| 8 | Place the COVIDISC in an oven at 65°C for ~ 45 min -1h |
| 9 | After returning to room temperature, read the fluorescence with the naked eye or a cell phone using appropriate filters |

### SUPPLEMENTARY MATERIAL 4

#### Experimental set-up to record fluorescence and signal processing

In the lab we used a dedicated set-up to acquire fluorescence, based on a Leica Z16 APO macroscope, equipped with FAM and Texas red cubes, and recorded images using an EM-CCD camera from Hamamatsu. This set up acquired an image of the reaction disks placed on a 65°C plate every 10 seconds.

Reaction disks were detected using active contours to automatically delineate their shape, using an image of the device taken before heating/starting the amplification. A single seed point inside each disk needs to be provided to perform each contour estimation. The Icy image analysis platform (www.icy.fr) was used to extract the data. The fluorescence signal plotted on Figure 2 is the average value of the 25% brighter pixels within the reaction disk, a simple approach to take into account profiles of high Ct (low viral load) samples, where the reaction happens and results in very bright pixels only in a subpart of the disk. We explored whether the heterogeneity of the fluorescence within the reaction disks could be used to reinforce the classification. We found out that the standard deviation (SD) of the fluorescence intensity within the reaction disk could increase the contrast between COVID-19 pools and negatives/other viruses samples (Figure S4). A SD threshold can thus classify all COVID-19 pools as positive and the negatives/non-specific samples as negative. We will explore in further experiments whether this parameter could complete/replace the fluorescence intensity to better classify tests as positive or negative.

**Figure S4.**
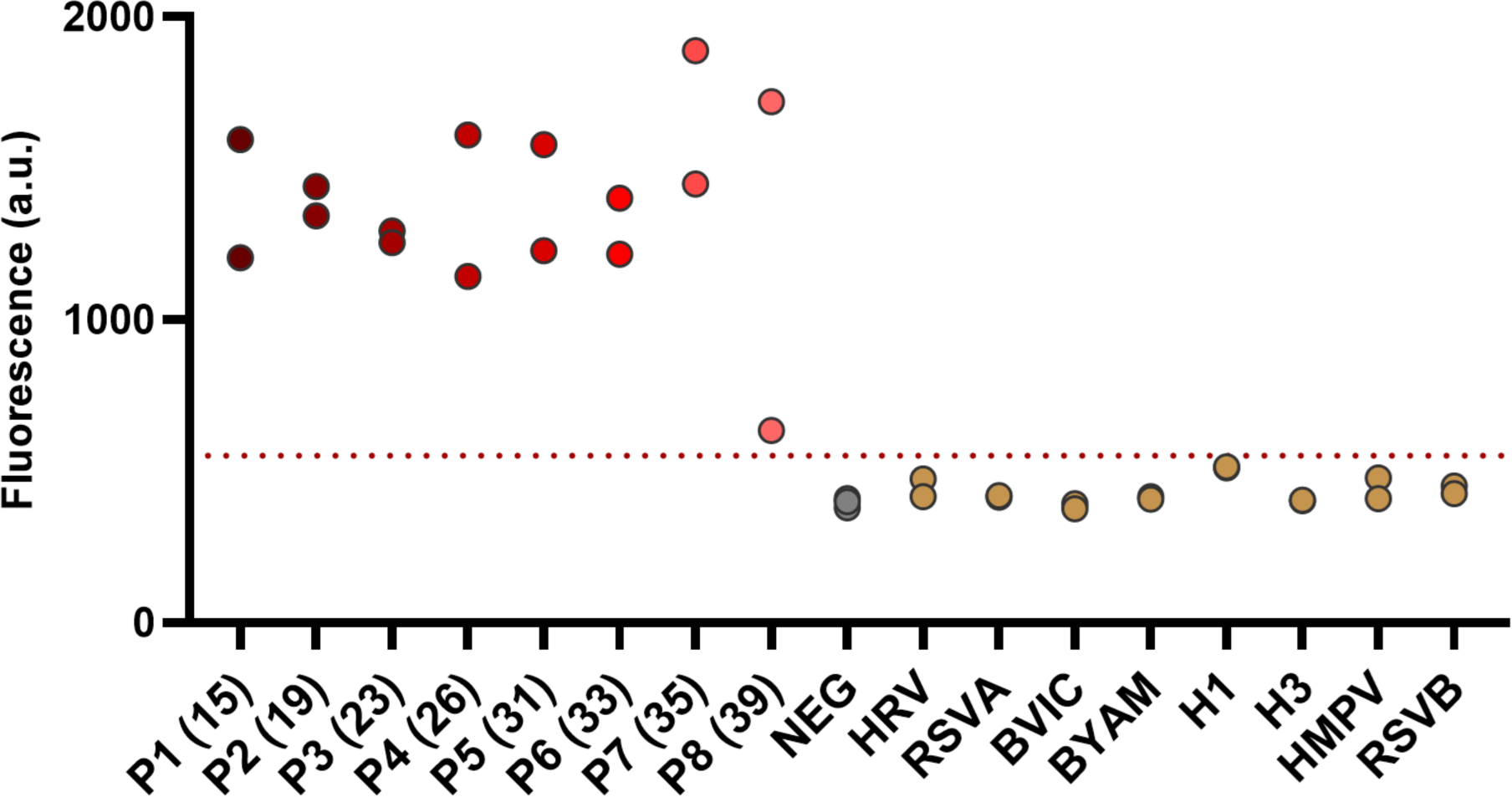
Sensitivity and Specificity measurements, using clinical COVID-19 and control samples. Same experiment and data as in Figure 2, where the signal is the standard deviation within the disk on the last (t=60min) picture. Red dotted line separating all positive from negative samples stands at SD = 550.

